# Identifying Active Paragonimiasis in Rural Nagaland in Northeastern India- a Community-Based Cross-Sectional Study

**DOI:** 10.64898/2026.07.02.26357096

**Authors:** Rohan Michael Ramesh, Kakhangchung Panmei, Azole, Arpitha Anbu Deborah, Thungbeni Humtsoe, Vinotsole Khamo, Sitara Swarna Rao Ajjampur, Abraham Joseph

**Author notes:** Co-corresponding author <u>Rohan Michael Ramesh</u>, The Wellcome Trust Research Laboratory, Division of Gastrointestinal Sciences, Christian Medical College, Vellore, Tamil Nadu, India-632004.

## Abstract

**Background:** Paragonimiasis is a neglected tropical disease (NTD) and of significant public health concern in northeastern India where it is frequently misdiagnosed as pulmonary tuberculosis. Transmission occurs through consumption of raw or undercooked freshwater crustaceans, a widespread dietary practice in this region. Accurate diagnosis will ensure cheap, effective treatment with praziquantel and avoid unnecessary prolonged tuberculosis therapy. This study determines the prevalence, specific risk factors for infection, and showcases a clinical scoring model to screen for active paragonimiasis in endemic areas.

**Methodology/Principal Findings:** A community-based cross-sectional survey was conducted in Medziphema block, Nagaland, in northeastern India screening 2,942 residents aged over 10 years. Participants reporting chronic cough (≥2 weeks) provided sputum and serum samples (93,3.16%). Sputum examination was screened for Paragonimus ova and acid-fast bacilli, and serum samples were tested for IgG antibodies against *Paragonimus heterotremus* using anti-Extra Secretory antigen by ELISA (positive case defined as titre ≥0.5 IU). IgG-confirmed paragonimiasis was detected in 30 individuals, yielding a population prevalence of 1.02% (30/2942) and 32.3% (30/93) amongst symptomatic participants. Sputum microscopy demonstrated limited sensitivity (3.3%), detecting cysts in only one confirmed case. Tuberculosis co-infection was documented in 6.7% of sero-positive cases. Multivariable regression identified significant associations with increasing age (adjusted (aOR) 1.04) previous tuberculosis history (aOR 4.62), monthly snail consumption frequency (aOR 1.05), ever smoking (aOR 3.51), and education beyond middle school (aOR 2.93). Haemoptysis (aOR 9.86) was the strongest symptomatic predictor. Clinical scoring models were developed, with haemoptysis consistently emerging as the most robust predictor across all models.

**Conclusions/Significance:** Pulmonary paragonimiasis constitutes a prevalent yet under-recognised condition in several southeast-Asian communities, necessitating systematic screening as a differential diagnosis for pulmonary tuberculosis. The clinical scoring model facilitates early case identification in resource-constrained settings. Public health interventions must address dietary practices, particularly consumption of raw or undercooked freshwater crustaceans, to interrupt transmission.

**AUTHOR SUMMARY:** “Paragonimiasis is a lung infection acquired by eating raw or undercooked crabs, prawns, or snails, a dietary practice in parts of Northeast India. Because its symptoms closely resemble pulmonary tuberculosis, many patients are misdiagnosed and receive prolonged TB treatment instead of a short course of praziquantel. We assessed how common active paragonimiasis is in rural Nagaland and identified factors linked to infection.

We surveyed 2,942 people in Medziphema Block and tested those with cough lasting at least 2 weeks. Active paragonimiasis was confirmed in 30 participants, giving a community prevalence of 1.02%, and in 32.3% of those with persistent cough. The disease was strongly linked to eating snails frequently, previous tuberculosis diagnosis (likely misdiagnosis), smoking, and older age. The standard sputum tests detected the infection in only 3% of cases.

A simple scoring system developed with just four questions, that health workers in remote clinics can use to identify people who need further testing.

These findings show that paragonimiasis is an important but overlooked cause of chronic cough in Nagaland. Simple clinical screening tools may help identify patients who need confirmatory testing. Public health efforts should promote safer preparation of freshwater crustaceans while respecting these deeply rooted cultural food traditions.”

## INTRODUCTION

Paragonimiasis is a food-borne trematodiasis of substantial yet under-recognised public health significance in Asia, Africa, and Latin America[1]. Caused by lung flukes of the genus *Paragonimus*, the infection afflicts an estimated 293 million people at risk globally, with several million infected, yet remains a neglected tropical disease in many endemic regions[2]. The most clinically important species include *Paragonimus westermani*, *Paragonimus heterotremus*, and *Paragonimus skrjabini*, with *P. heterotremus* particularly prevalent across Southeast Asia and the Indian subcontinent[3]. The lifecycle of *Paragonimus* species involves freshwater snails and crustaceans (crabs, crayfish, and prawns) as intermediate hosts, with humans as the definitive host. Human infection occurs through consumption of raw or inadequately cooked crustaceans harbouring metacercariae, which penetrate the intestinal wall, migrate through the peritoneal cavity and diaphragm, and mature into adult flukes in the lung parenchyma. The incubation period ranges from two days to two months[4].

Clinically, paragonimiasis presents predominantly as pulmonary disease (76–90% of cases), with extrapulmonary manifestations, cerebral, abdominal, cutaneous, and cardiovascular (2–10% of patients). Pulmonary paragonimiasis is characterised by chronic cough, haemoptysis, dyspnoea, chest pain, and constitutional symptoms including fever, weight loss, and night sweats [5]. Notably, up to 6% of infected individuals may remain asymptomatic [6]. The clinical and radiological features of pulmonary paragonimiasis bear striking resemblance to pulmonary tuberculosis, leading to frequent misdiagnosis and inappropriate treatment with anti-tubercular regimens in tuberculosis-endemic regions[7,8]. This diagnostic confusion has substantial implications. Treatment with praziquantel (75 mg/kg/day over three days) is simple, effective, and costs approximately 250 rupees, whereas misdiagnosis as tuberculosis subjects patients to over six months of therapy at considerably higher cost[9,10]. In regions where both infections coexist, dual infection may be misinterpreted as tuberculosis treatment failure. Early and accurate diagnosis is therefore essential to prevent unnecessary treatment burden and ensure appropriate intervention [3].

In Northeast India, paragonimiasis represents an important but under-studied public health concern. The endemic status has been documented in multiple states including Manipur, Assam, Arunachal Pradesh, and Nagaland, driven largely by dietary practices involving consumption of raw or undercooked freshwater crustaceans, a deeply ingrained cultural tradition across diverse tribal communities [7,11–14]. In Nagaland, facility based investigations identified paragonimiasis among patients presenting to tuberculosis clinics, with varied detection rates of 50% (7/14) in Phek District and 3·1% (3/96) in Mon District [7,15]. However, community-based data on population-level prevalence and epidemiological determinants of active infection remain limited.

Against this backdrop, we conducted a community-based cross-sectional study in Medziphema Block, Dimapur District, Nagaland, a region characterised by extensive paddy cultivation, perennial rivers, and widespread consumption of freshwater crustaceans, to determine the prevalence, identify specific risk factors for infection, and develop a clinical scoring model to screen for active paragonimiasis. This study intends to address critical knowledge gaps regarding the burden of paragonimiasis in Nagaland, inform targeted public health interventions, and develop evidence-based strategies to facilitate early case identification in resource-limited primary care settings where paragonimiasis and tuberculosis overlap.

## METHODS

### Study Design

A community-based, cross-sectional survey was conducted in Medziphema Block, Chümoukedima District (formerly Dimapur District), in Nagaland, northeastern India between November 2015 and October 2016. Situated 44 km from Kohima and 33 km from Dimapur, the block spans a transitional zone between the hills and the plains and encompasses Medziphema town and 21 surrounding villages. Covering 388 km², it accommodates approximately 24,149 residents at a density of 62 persons/km². Nearly 76.6% of inhabitants belong to various Naga tribes classified as Scheduled Tribes under Article 342 of the Indian Constitution [16,17]. Health care in Medziphema Block is provided by one community health centre in Medziphema town, five primary health centres, and three subcentres. While most villages occupy hilly terrain, those nearer the town lie on plains dominated by paddy fields irrigated by multiple streams and the Dzumha and Chate rivers[18]. (**Fig 1**)

**Fig 1.**
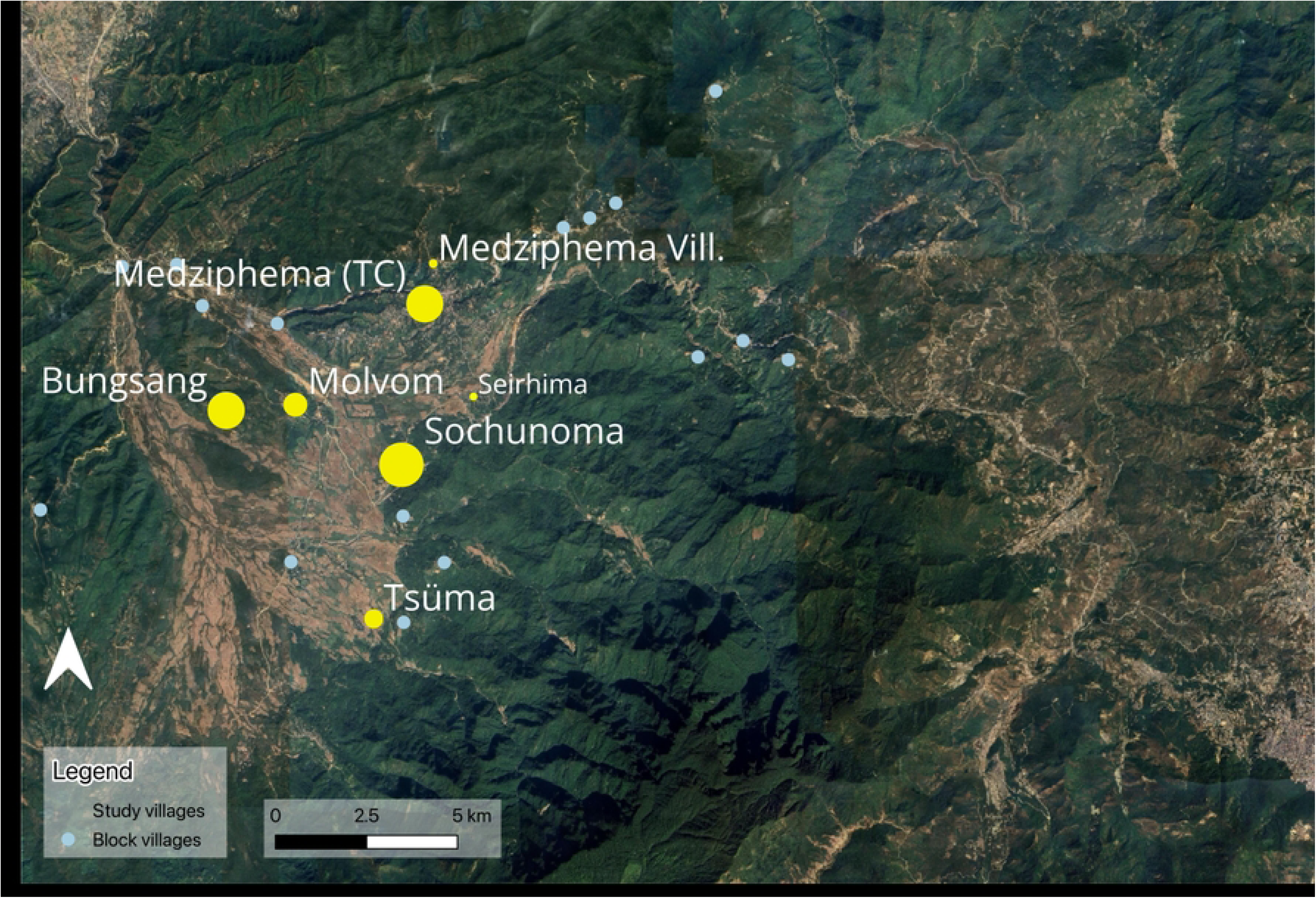
Map of the study sites in Medziphema block, Nagaland

### Sample Size and Sampling

The required sample size was estimated to detect a 6·7% prevalence of active paragonimiasis[19] with 20% relative precision at the 95% confidence level. Incorporating a design effect of 2·0 and 10% loss to follow-up, we calculated that 2942 individuals would need to be enrolled. A cluster sampling strategy was utilized with one urban cluster (Medziphema town) and six rural clusters (Medziphema village, Bunsong, Molvom, Sirii Angami, Suchunoma, and Tsuuma) selected in the block.

### Inclusion and Exclusion Criteria

Participants aged ≥10 years who had resided in Medziphema Block for more than two years and provided informed consent were eligible; those who declined consent or were absent during initial household visits were excluded.

### Community Engagement and Recruitment

Village leaders were approached and provided with an information sheet regarding the study. Community Volunteers (CVs), recommended by local Village Council, was recruited and received intensive training focusing on the informed consent process, administration of the study questionnaire, identification of symptoms, and practical sessions with mock drills. The CV conducted household visits, obtained informed consent, and administered a structured questionnaire. The survey documented sociodemographic characteristics, health service utilization, dietary habits and symptoms indicative of paragonimiasis and tuberculosis.

### Data Collection Tools and Procedures

Data were collected using a purpose-designed, tablet-based questionnaire, which was piloted for validity and reliability before deployment. Information captured comprised: sociodemographic characteristics (age, sex, education, occupation, household composition); health-service use; detailed dietary history with emphasis on consumption and preparation of freshwater crustaceans (snails, crabs, prawns); and clinical manifestations, notably chronic cough (≥2 weeks), haemoptysis, chest or abdominal pain, fever, and malaise.

### Case Identification and Sample Collection

Participants reporting a cough of ≥2 weeks were counselled and provided additional verbal and written consent for specimen collection. Three sputum specimens were obtained per participant (spot on day 1, early-morning on day 2 and day 3), stored at 2–8 °C in insulated transport boxes and delivered to the Christian Institute of Health Sciences and Research (CIHSR), Dimapur laboratory located 33 kms away for microscopy. On day 3, 4 mL of venous blood was drawn under aseptic conditions; serum was separated by centrifugation, frozen at –80 °C, and transported weekly to the Regional Medical Research Centre (RMRC), Dibrugarh, for ELISA.

### Laboratory Methods

For examination of Acid-Fast Bacilli, the standard Ziehl–Neelsen staining was performed according to National TB Elimination Program – NTEP guidelines. For the detection of Paragonimus ova, concentration method, was performed using Formal Ether Concentration Technique (FECT). For the FECT, sputum was homogenized with 1% NaCl, 10% formalin added, mixed and centrifuged. Supernatant was discarded and the sediment was examined for direct examination. Wet mount was performed for direct detection observed under 10x objective (100x magnification) and 40x objective (400x magnification) [20]. For serology, an anti–excretory–secretory antigen IgG ELISA validated at RMRC Dibrugarh, with 100% sensitivity and specificity for *Paragonimus heterotremus*, was used; titres >0.5 IU (ELISA OD 0.494, the ROC-derived optimal cutoff with 100% sensitivity and specificity) or identification of ova in sputum confirmed infection [21].

### Clinical Management and Referral

Confirmed Paragonimiasis cases received oral praziquantel (75mg/kg body weight for three consecutive days). Participants diagnosed with tuberculosis were referred to the nearest DOTS centre for anti-tubercular therapy in accordance with RNTCP guidelines. In this study, ‘active paragonimiasis’ was defined as laboratory-confirmed *Paragonimus* infection (Paragonimus eggs detected in sputum or serum IgG ELISA titre ≥0.5 IU against *Paragonimus heterotremus* ES-antigen) among community participants with cough lasting at least two weeks.

### Quality Assurance and Data Management

Questionnaire instruments were pilot-tested and revised following community focus group discussions. CVs’ data collection practices were supervised by the field supervisor, with periodic quality checks. Errors were resolved by verification of source documentation. All samples were handled using standardized cold chain procedures.

### Statistical Analysis

Descriptive statistics summarized demographic, clinical and laboratory data. Prevalence rates were calculated with 95% confidence intervals. Univariate analysis (chi-square) identified factors potentially associated with paragonimiasis at *p < 0.1*. The variables meeting this threshold or reported as established risk factors in prior studies such as age, gender, education previously diagnosed as TB, consumption of snails, prawns or crabs and smoking status were entered into multivariable logistic regression models to identify independent associations with confirmed disease. Results are reported as adjusted odds ratios (aORs) and 95% CIs and the most fitting model including age, gender, education, past history of TB, smoking and monthly consumption of snails was selected. In a sensitivity analysis, we examined the effect of inclusion of monthly consumption of crabs and prawns as additional candidate predictors.

Four predictive clinical scoring models were developed by translating adjusted odds ratios (ORs) into weighted point values. The performance of each model was assessed by calculating the area under the receiver-operating-characteristic curve (ROC-AUC) and comparing results to determine the optimal model. To ensure ease of use in routine clinical settings, only the most strongly predictive and common patient history variables were retained in the final scoring system.

### Ethical Considerations

The study protocol was approved by the Institutional review board of CIHSR, Dimapur, Nagaland. Written informed consent was obtained from all participants, with additional procedures in place to protect privacy, minimize risks, and ensure linkage to care for affected individuals.

## RESULTS

Overall, 2 942 individuals aged 10 years or older who had resided in Medziphema Block for at least two years were enrolled in the study. The median age was 32 years (IQR 21-47), with the largest proportion (22.7%) in the 20-29 years age group. Women comprised 57.0% of participants. Most participants resided in urban wards of Medziphema town (73.3%), with the remainder in rural villages. The cohort was ethnically diverse: 86.1% belonged to tribal communities, predominantly Angami (41.0%), Kuki (20.1%), and Nepali (9.8%). The overall, educational status was commendeble, with 40.8% having completed at least secondary school, and the mean household size was six members. Dietary patterns reflected the cultural context: 99.5% of participants consumed non-vegetarian diets, with chicken (97.5%), fish (95.1%), and pork (94.6%) most commonly eaten. Notably, consumption of freshwater organisms implicated in paragonimiasis transmission was widespread: snails (74.7%), crabs (63.8%), frogs (62.7%), and prawns (60.8%). Current smoking was reported by 13.3% of participants, while 7.7% were former smokers **(S1 Table)**

### Prevalence of Active Paragonimiasis

Of these participants, 93 (3.2%) reported a cough lasting two weeks or more and provided three sputum specimens alongside a serum sample for diagnostic evaluation. Serological testing using IgG ELISA identified 30 individuals with titres ≥0.5, corresponding to an overall community prevalence of 1.02% (30/2942) and a prevalence of 32.3% among symptomatic participants (30/93).

### Detection Rate of Sputum Microscopy

Of the 93 symptomatic individuals, only one demonstrated Paragonimus ova in sputum microscopy, corresponding to a sensitivity of 3.30% (1/30) among ELISA-confirmed cases; specificity was 100%.

### Tuberculosis and Co-infection

Among participants with chronic cough, the prevalence of sputum-positive tuberculosis was 2.15% (2/93). Co-infection with paragonimiasis occurred in 6.67% (2/30) of ELISA-confirmed cases.

### Risk Factors for Active Pulmonary Paragonimiasis

In multivariable logistic regression analysis, the final model revealed several factors independently associated with active paragonimiasis. Age was independently associated with paragonimiasis risk, with odds increasing by 4% for each additional year of age (adjusted OR 1.04, 95% CI 1.02-1.06). Prior history of tuberculosis was associated with over fourfold increased odds (adjusted OR 4.62, 95% CI 1.79-11.94) and past smoking (adjusted OR 3.51, 95% CI 1.13-10.94) also conferred higher risk. Dietary exposures significantly associated with infection included monthly consumption of snails (adjusted OR 1.05, 95% CI 1.01-1.08). Higher education (≥secondary school) was also independently associated (adjusted OR 2.93, 95% CI 1.12-7.67) with active paragonimiasis (**Table 1**). Sensitivity analysis with monthly consumption of prawns and crabs as additional candidate predictors to the final selected model attenuated the association with all the other variables. Multiple combinations of including consumption of snail, prawns and crabs demonstrated collinearity among these three variables, as indicated in **(S2 Table).**

**Table 1.**
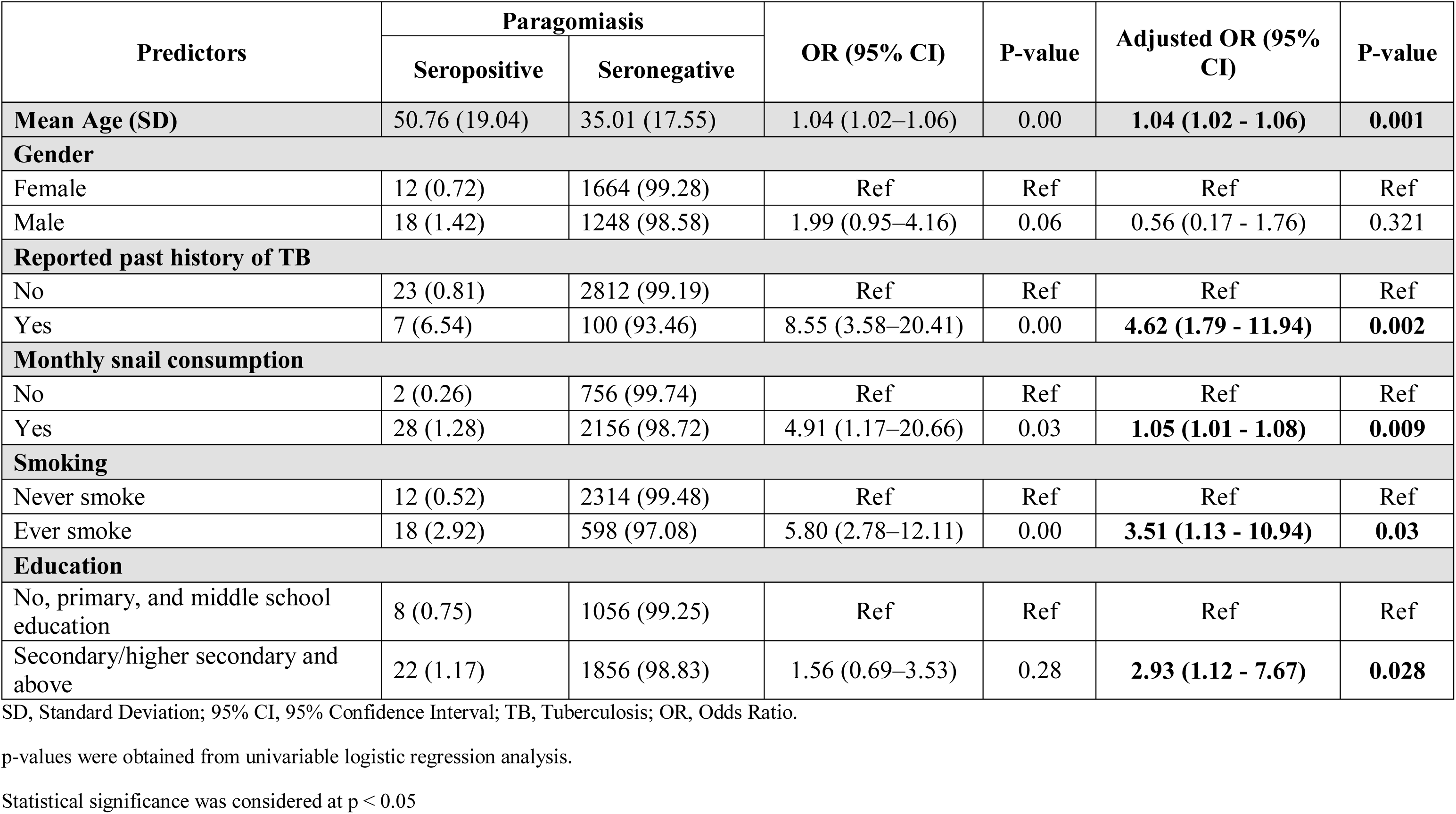
Comparison of Baseline Characteristics Between the Seropositive and Seronegative Groups.

### Clinical score models for endemic sites

Four clinical scoring models were developed to diagnose active paragonimiasis in the community, each differing with selection and weighting of the included variables. The models included variables such as prior tuberculosis diagnosis, smoking history, monthly snail consumption, fever and presence of blood in sputum. Although abdominal pain demonstrated a statistically significant association with paragonimiasis (aOR 3.46, 95%CI 1.31–9.16), it was not included due to the absence of a biologically plausible explanation and instead fever, a more objective symptom that also overlaps with TB was selected. ROC curve was generated to compare the scoring models (**Fig 2**). Model 1 demonstrated the highest discriminatory performance with an ROC-AUC of 0.87, followed closely by Model 2 with 0.85. Models 3 and 4, while simpler, showed modestly lower AUCs of 0.81 and 0.79 respectively. Given the aim for a practical, easy-to-use tool in low-resource community settings, Model 2 strikes the best balance between accuracy and usability, omitting history of fever, making it an optimal scoring system for early identification and management of active paragonimiasis in endemic areas (**Table 2**). However, blood in sputum emerged as the strongest independent symptomatic predictor (aOR 9.86, 95%CI 2.11–46.09), and could potentially serve as a critical clinical trigger for the investigation of pulmonary paragonimiasis (S3 Table).

**Fig 2.**
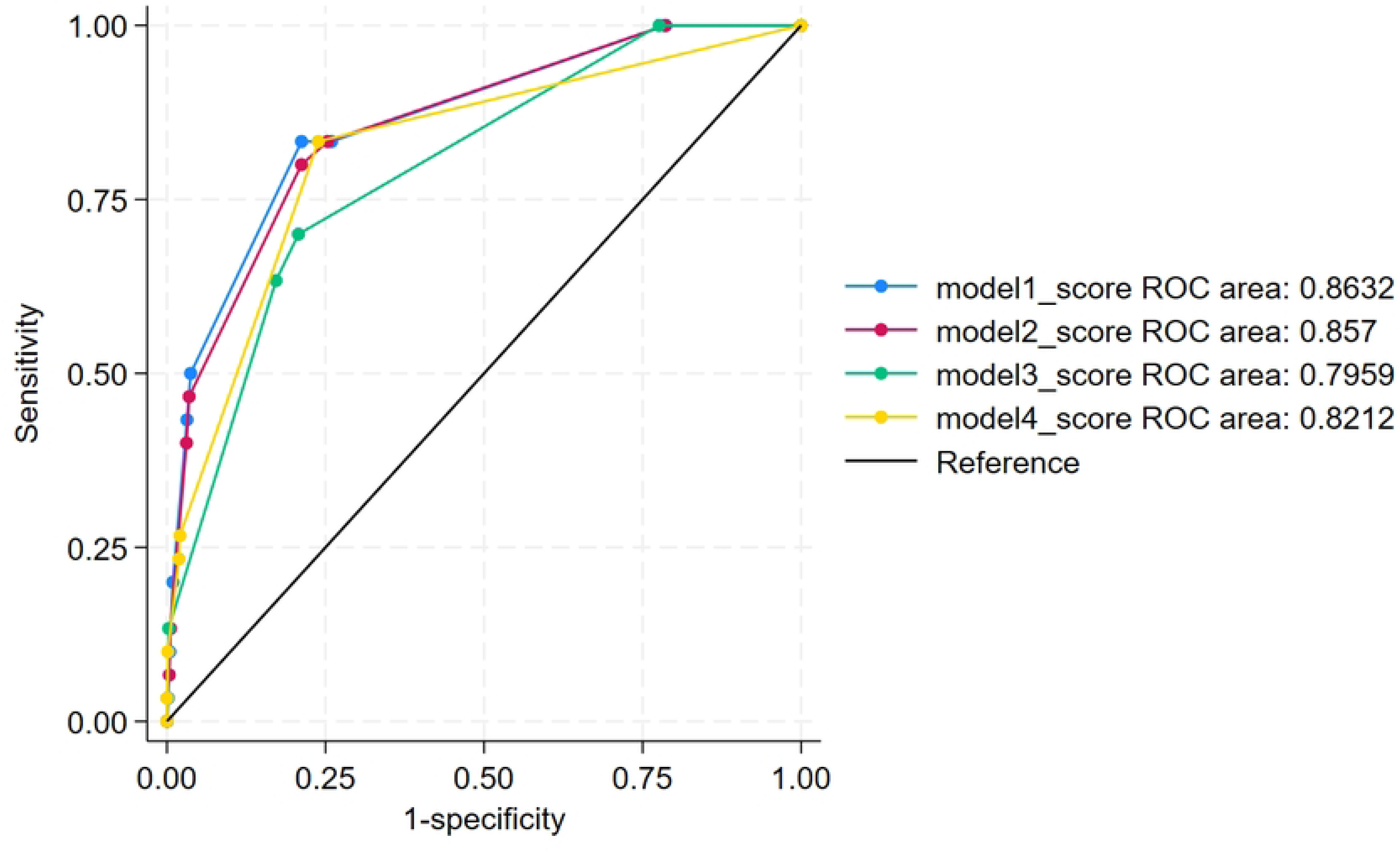
Receiver operating characteristics curve comparing the various clinical score models

**Table 2.**
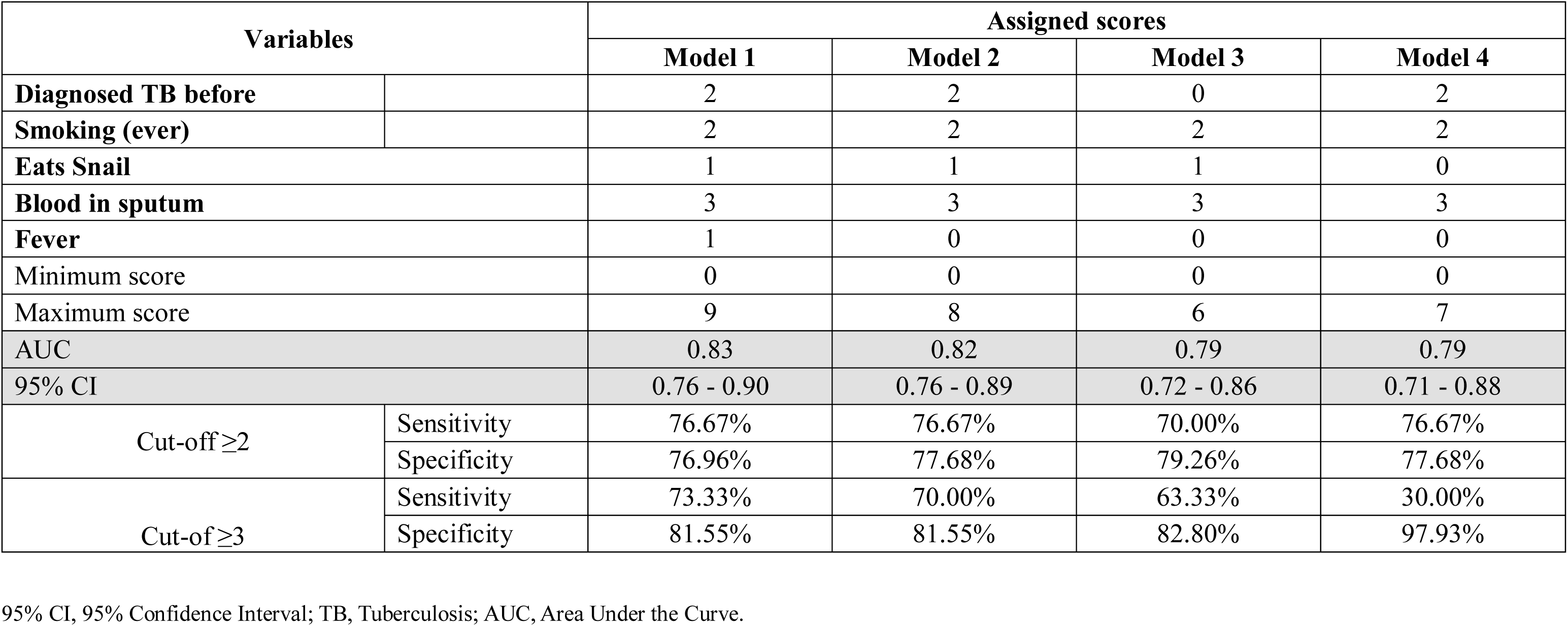
Assigned Scores for Predictor Variables Across the Five Risk Prediction Models.

## DISCUSSION

This community-based cross-sectional study reveals that active paragonimiasis remains a substantial yet under-recognised public health burden in Nagaland, Northeast India, with a seroprevalence of 1.02% in the general population and 32.3% among individuals presenting with chronic cough, positioning Medziphema Block as an endemic area. Prevalence estimates across Northeast India range from 2% to 36%, though our community-wide figure is notably lower than previously documented rates in tribal populations of Arunachal Pradesh (3.7 to 8.2%) but comparable to surveys in Assam [12,13,22].

The high prevalence among symptomatic participants underscores a critical diagnostic challenge: nearly one in three individuals with chronic cough harboured paragonimiasis rather than tuberculosis. The remarkably low sensitivity of sputum microscopy (3.3%), considerably lower than other reports from facility-based studies, demonstrates that conventional parasitological diagnosis is inadequate for community-based case detection[23,24]. Several factors likely contributed to the low sputum microscopy yield: collection of only two early morning samples (rather than consecutive daily samples), prolonged transport time causing egg degradation, and the possibility that some seropositive participants were in early infection stages before egg production commences at 2-3 months post-infection[4]. ELISA seems to be the most likely choice for mass screening in endemic settings with a high sensitivity and specificity, decreasing the possibility of cross reactivity with other parasitic infections [21]. However, limitations include antibody persistence for 4-18 months post-treatment and species-specificity to *P. heterotremus*, potentially missing other Paragonimus species. A locally developed rapid diagnostic test kits utilising species-specific antigens from Paragonimus endemic to Nagaland offers a promising tool for point-of-care diagnosis in resource-limited primary health facilities [25]

Tuberculosis co-infection occurred in 6.67% of paragonimiasis cases, with 2.15% of symptomatic participants having TB. This overlap, as previously documented by Singh and Das et al in Nagaland, complicates diagnosis and confounds treatment in resource-limited settings where the burden of diseases are high [8,15,26]. Educating primary care health workers and clinicians in health facilities including TB clinics to include paragonimiasis in the differential diagnosis of smear-negative chronic respiratory illness could reduce diagnostic delays and improve treatment outcomes in endemic areas.

Multivariable analysis revealed that prior tuberculosis history was the strongest predictor of paragonimiasis (adjusted OR 4.62, 95% CI 1.79-11.94). This association likely reflects diagnostic confusion rather than biological causation, as both diseases share similar respiratory symptoms such as chronic cough, blood in sputum, and chest pain[4]. Many participants diagnosed with tuberculosis may have had unrecognised paragonimiasis, especially given the rarity of parasitological confirmation in primary care [26]. Alternatively, pre-existing tuberculous lung damage may create a more permissive environment for parasite establishment, though less plausible. Past smoking history (adjusted OR 3.51, 95% CI 1.13-10.94) similarly conferred elevated risk, plausibly through compromised respiratory epithelium and impaired mucociliary clearance mechanisms that may facilitate parasite penetration and establishment. Dietary exposures demonstrated the expected associations with transmission: snail consumption showed a modest but statistically significant association (adjusted OR 1.05 per consumption occasion, 95% CI 1.01-1.08), consistent with snails serving as the first intermediate host in the Paragonimus life cycle. Each additional year of age independently increased the odds of paragonimiasis by 4% (adjusted OR 1.04, 95% CI 1.02-1.06), consistent with cumulative dietary exposure over the lifespan. The unexpected positive association between higher education (adjusted OR 2.93, 95% CI 1.12-7.67) and paragonimiasis contradicts conventional patterns observed with neglected tropical diseases. This may reflect unmeasured confounding related to dietary preferences related to affordability, or increased frequency of intake among educated populations in this specific context [1]. Collectively, these findings emphasise that transmission is sustained primarily through culturally embedded dietary practices involving raw or inadequately cooked freshwater crustaceans, and that effective prevention strategies must address both food safety education and the deeply rooted culinary traditions of this community.

The development of clinical scoring models represents a pragmatic response to the diagnostic challenges identified in this study, offering a potentially transformative tool for case detection in resource-limited primary care settings where serological testing remains inaccessible. Model 2, which balances diagnostic accuracy with practical implementation feasibility, represents the optimal choice for field deployment in community screening programs, particularly given its reliance on simple clinical history and physical examination findings that require neither laboratory infrastructure nor specialized training. A simple diagnostic checklist incorporating history of prior tuberculosis, smoking status, monthly snail consumption, and blood in sputum may be employed during initial clinical assessment of patients presenting with persistent cough (≥2 weeks’ duration) in resource-limited primary care settings. Implementation of this screening tool could facilitate early detection and increase clinical suspicion for paragonimiasis in populations where conventional diagnostic facilities are unavailable. Among these predictors, blood in sputum emerged as the strongest independent symptomatic indicator, underscoring the potential utility of bloody sputum as a critical clinical trigger for investigating paragonimiasis in endemic areas. However, these models require external validation in other endemic populations with differing prevalence rates and risk factor profiles before widespread implementation can be recommended. The clinical scoring approach addresses a fundamental gap in paragonimiasis control: the absence of affordable, point-of-care diagnostic tools suitable for primary health facilities where the majority of at-risk populations first present with respiratory symptoms. By enabling front-line health workers to identify high-probability cases warranting further investigation or empirical treatment, these models could substantially reduce diagnostic delays, prevent unnecessary tuberculosis treatment, and improve health outcomes in endemic communities where paragonimiasis remains persistently under-recognized despite its considerable disease burden.

Effective control of paragonimiasis in endemic areas requires a comprehensive strategy integrating treatment, prevention, surveillance, and environmental interventions. Praziquantel offers highly effective treatment with cure rates exceeding 90%, and is substantially more cost-effective than the prolonged multi-drug regimens required for tuberculosis. Prevention efforts must prioritise culturally sensitive health education targeting high-risk populations, particularly rural communities where consumption of raw or inadequately cooked freshwater crustaceans is deeply embedded in traditional dietary practices. Successful control programmes in Arunachal Pradesh demonstrate that community education combined with mass treatment can significantly reduce prevalence when implemented with cultural sensitivity and sustained engagement[12]. Surveillance strategies should leverage the clinical scoring models developed in this study to enable targeted case detection in primary care settings, with integration into existing tuberculosis screening programmes to maximise efficiency and reduce duplication[27]. Longer-term control will require environmental interventions addressing the parasite’s complex life cycle, including improved water and sanitation infrastructure, although the feasibility of controlling intermediate hosts (snails and crustaceans) in natural freshwater ecosystems remains limited. Post-treatment monitoring through repeat serological testing and symptom assessment should be implemented to confirm cure and identify treatment failures, particularly in areas where reinfection risk remains high due to persistent dietary practices. These multifaceted interventions, adapted to local epidemiological and cultural contexts, offer the most promising pathway toward reducing the substantial but neglected burden of paragonimiasis in Northeast India.

Key strengths of this study include the large community-based design (2,942 participants), validated serological testing, comprehensive multivariable risk factor analysis, development of clinical scoring models, and assessment of tuberculosis co-infection in this dual-endemic population. Few limitations of this study include the cross-sectional design precluding causal inference; serological testing potentially detecting past infection and being specific to *P. heterotremus*; single-timepoint sputum collection missing intermittent shedding; recall bias in dietary reporting; and geographic restriction to one block limiting generalisability. The clinical scoring models require external validation in diverse settings before widespread implementation. These constraints should be considered when interpreting findings and planning future research.

## Data Availability

The de-identified data that support the findings of this study are available from the corresponding author upon reasonable request.

## ACKNOWLEDGMENTS

We acknowledge the Indian Council of Medical Research for funding this study. We are grateful to the Regional Medical Research Centre, North-East Region, Dibrugarh, Assam, and Dr Rajguru for conducting serological assays and providing technical expertise. We thank the Village Councils and community leaders of Medziphema Block for their permission and support, and the laboratory staff at Christian Institute of Health Sciences and Research, Dimapur, for specimen processing. Above all, we are deeply indebted to the 2,942 participants from Medziphema Block who generously volunteered their time and provided specimens. Their participation made this research possible and will contribute to improved health outcomes for paragonimiasis-affected communities in Northeast India. We also thank Dr Karthik Gunasekaran for his guidance in developing the clinical score models.

## Supporting Information

**S1 Table**. Baseline demographic, dietary and smoking history

**S2 Table**. Comparison of multivariable regression models

**S3 Table**. Symptoms predicting pulmonary paragonimiasis in Nagaland

